# Histology-based Prediction of Therapy Response to Neoadjuvant Chemotherapy for Esophageal and Esophagogastric Junction Adenocarcinomas Using Deep Learning

**DOI:** 10.1101/2023.03.01.23286553

**Authors:** Fabian Hörst, Saskia Ting, Sven-Thorsten Liffers, Kelsey L. Pomykala, Katja Steiger, Markus Albertsmeier, Martin K. Angele, Sylvie Lorenzen, Michael Quante, Wilko Weichert, Jan Egger, Jens T. Siveke, Jens Kleesiek

**Affiliations:** Institute for Artificial Intelligence in Medicine (IKIM), University Hospital Essen (AöR), Essen, Germany; Cancer Research Center Cologne Essen (CCCE), West German Cancer Center Essen, University Hospital Essen (AöR), Essen, Germany; Institute of Pathology Nordhessen, Kassel, Germany; Bridge Institute of Experimental Tumor Therapy, West German Cancer Center Essen, University Hospital Essen (AöR), Essen, Germany; Division of Solid Tumor Translational Oncology, German Cancer Consortium (DKTK, Partner site Essen) and German Cancer Research Center (DKFZ), Heidelberg, Germany; Institute of Pathology, Technical University of Munich (TUM), Munich, Germany; Department of General, Visceral and Transplantation Surgery, LMU University Hospital, Ludwig-Maximilians-Universität (LMU), Munich, Germany; Clinic for Internal Medicine III, University Hospital rechts der Isar, Technical University of Munich (TUM), Munich, Germany; Clinic for Internal Medicine II, Gastrointestinal Oncology, University Medical Center of Freiburg, Freiburg, Germany; Department of Internal Medicine II, University Hospital rechts der Isar, Technical University of Munich (TUM), Munich, Germany; German Cancer Consortium (DKTK), Heidelberg, Germany; German Cancer Research Center (DKFZ), Heidelberg, Germany; West German Cancer Center, Department of Medical Oncology, University Hospital Essen (AöR), Essen, Germany; Medical Faculty, University Duisburg-Essen, Essen, Germany; German Cancer Consortium (DKTK, Partner site Essen), Heidelberg, Germany

## Abstract

**Background:** Quantifying treatment response to gastroesophageal junction (GEJ) adenocarcinomas is crucial to provide optimal therapeutic strategy. Routinely taken tissue samples provide an opportunity to enhance existing PET/CT-based therapy response evaluation. Our objective was to investigate if deep learning algorithms are capable to predict the therapy response of GEJ patients to neoadjuvant chemotherapy based on histological tissue samples.

**Methods:** This diagnostic study recruited 67 patients with GEJ I-III from the multicentric non-randomized MEMORI trial including 3 German university hospitals TUM (Munich), LMU (Munich), and UME (Essen). All patients underwent baseline PET/CT scans and esophageal biopsy before and 14-21 days after treatment initiation. Treatment response was defined as a ≥ 35% decrease in SUVmax from baseline. Several deep learning algorithms were developed to predict PET/CT-based responders and non-responders to neoadjuvant chemotherapy using digitized histopathological whole slide images.

**Results:** The resulting models were trained on TUM (n=25 pre-therapy, n=47 on-therapy) patients and evaluated on our internal validation cohort from LMU and UME (n=17 pre-therapy, n=15 on-therapy). Compared with multiple architectures, the best pre-therapy network achieves an area under the precision-recall curve (AUPRC) of 0.81 (95% confidence interval (CI), 0.61-1.00), area under the precision-recall curve (AUPRC) of 0.82 (95% CI, 0.61-1.00), balanced accuracy of 0.78 (95% CI, 0.60-0.94), and a Matthews correlation coefficient (MCC) of 0.55 (95% CI, 0.18-0.88). The best on-therapy network achieves an AUROC of 0.84 (95% CI, 0.64-1.00), AUPRC of 0.82 (95% CI, 0.56-1.00), balanced accuracy of 0.80 (95% CI, 0.63-1.00), and MCC of 0.71 (95% CI, 0.38-1.00), solving a task beyond the pathologists’ capabilities.

**Conclusions:** The findings suggest that the networks can predict treatment response using WSI with high accuracy even pre-therapy, suggesting morphological tissue biomarkers. Subject to further validation, this could lead to earlier therapy intensification compared to current PET/CT diagnostic system for non-responder.

## Introduction

The incidence of gastroesophageal junction (GEJ) adenocarcinomas has rapidly increased over the last 30 years, especially in Europe, North America, and Australia.^1–3^ Although the prognosis has improved over time, it remains unfavourable with only 20% of patients in Western populations surviving 5 years.^1,4–6^

In locally advanced GEJ, perioperative chemotherapy (e.g., FLOT4^7^) and neoadjuvant radiochemotherapy (e.g., CROSS^8^) strategies have successfully improved survival. However, they have not been compared to each other, thus leaving the best treatment modality unknown so far. Several trials have investigated early PET imaging for therapy response prediction. We recently performed the MEMORI trial (NCT02287129) which evaluated PET-directed neoadjuvant chemotherapy (CTX) or salvage chemoradiotherapy (CRT). Importantly, we assembled sequential, high-quality tumor biopsies at FDG-PET/CT imaging time points pre-therapy and during therapy for GEJ.^9^ This trial demonstrated an improved rate of negative surgical margins and pathologic complete remission when patients underwent salvage intensified CRT after not responding to standard neoadjuvant CTX determined by PET response 14-21 days after chemotherapy initiation.^9^ Thus, early treatment response assessment can inform and improve patient care in this setting.^9,10^

However, acquiring PET/CT images pre- and during therapy is logistically demanding, cost-intensive, and the patient is exposed to radioactive tracers twice early during treatment. Furthermore, at least 14 days are needed to determine responder status based on metabolic uptake. On the contrary, biopsy samples are routinely taken for diagnosis prior to therapy. Nonetheless, trying to determine the effect of neoadjuvant CTX solely based on hematoxylin and eosin (H&E) stained biopsy samples still needs to be solved, as no predictive biomarkers nor histological patterns for this are known. In this study, we analyzed tumor features of GEJ carcinomas in digitized histology images pre- and during treatment by utilizing deep learning (DL) approaches to predict the treatment response to neoadjuvant CTX, focusing on the routinely sampled pre-therapy biopsy slides.

For the computational assessment, tissue slides are scanned at high resolution resulting in whole slide images (WSI). Some challenges associated with WSI are the large image size, high morphological variance, inconsistent staining, and information at different magnifications (local vs. global structure).^11,12^ Despite these challenges, DL approaches have been successfully applied to various low-level image analysis tasks, including image preprocessing^13–15^, disease classification^16^, cell detection^17–19^, and segmentation^20,21^, as well as higher-level tasks such as predicting disease prognosis and diagnosis^22^, patient survival analysis^23^, treatment prognosis^24,25^, or identification of complex biomarkers like microsatellite instability detection^26^. Especially for high-level tasks, the mapping between tissue morphology and the global label is often unknown, notably, if pathologists have no prior knowledge and identifying the underlying relationship itself is part of the research.^27^

Thus, to predict the treatment response to neoadjuvant CTX, we used algorithms relying on weak supervision without any tissue annotations. All algorithms were validated on an internal validation cohort.

## Materials and Methods

The MEMORI study was approved by the Ethics Committee of University Hospital Rechts der Isar, Munich, Germany, and has been reported previously^9^. This study was carried out in accordance with the Declaration of Helsinki. We followed the Transparent Reporting of a Multivariable Prediction Model for Individual Prognosis or Diagnosis (TRIPOD) reporting guideline.^28^

### Patients and Dataset

A patient cohort from the non-randomized MEMORI trial with histologically confirmed GEJ I-III according to Siewert classification^29^ from three German university hospitals (Munich, TUM; Munich, LMU; Essen, UME), treated between December 1, 2014 and December 31, 2018, was used in this study. Patients were excluded if they had previous radiotherapy targeted at the thorax, existing distant metastases (M1b), or tumor infiltration into the tracheobronchial system (see supplementary Table S1 for all criteria).

All patients initially underwent baseline 18F-FDG PET/CT and tumor biopsy followed by one cycle of chemotherapy (pre-therapy, d1). PET/CT and endoscopic esophageal biopsies were repeated on days 14-21 (on-therapy, d14-21) after the first PET/CT scan. Based on metabolic tumor activity quantified by the PET standardized uptake value (SUV), patients with a ≥ 35 % decrease in d14-21 SUVmax compared to d1 baseline were defined as responders (R), otherwise as non-responders (NR), similar to the previous MUNICON I+II trials^10,30–32^. Responder patients continued with CTX prior to surgery, non-responders switched to salvage chemoradiotherapy. Interventions conducted after the second PET/CT scan to determine the treatment response of the first CTX cycle on d14-d21 were not considered in this investigation. A flowchart depicting the process of patient enrollment and subgroup formation with intervention events is provided in Figure 1.

**Figure 1.**
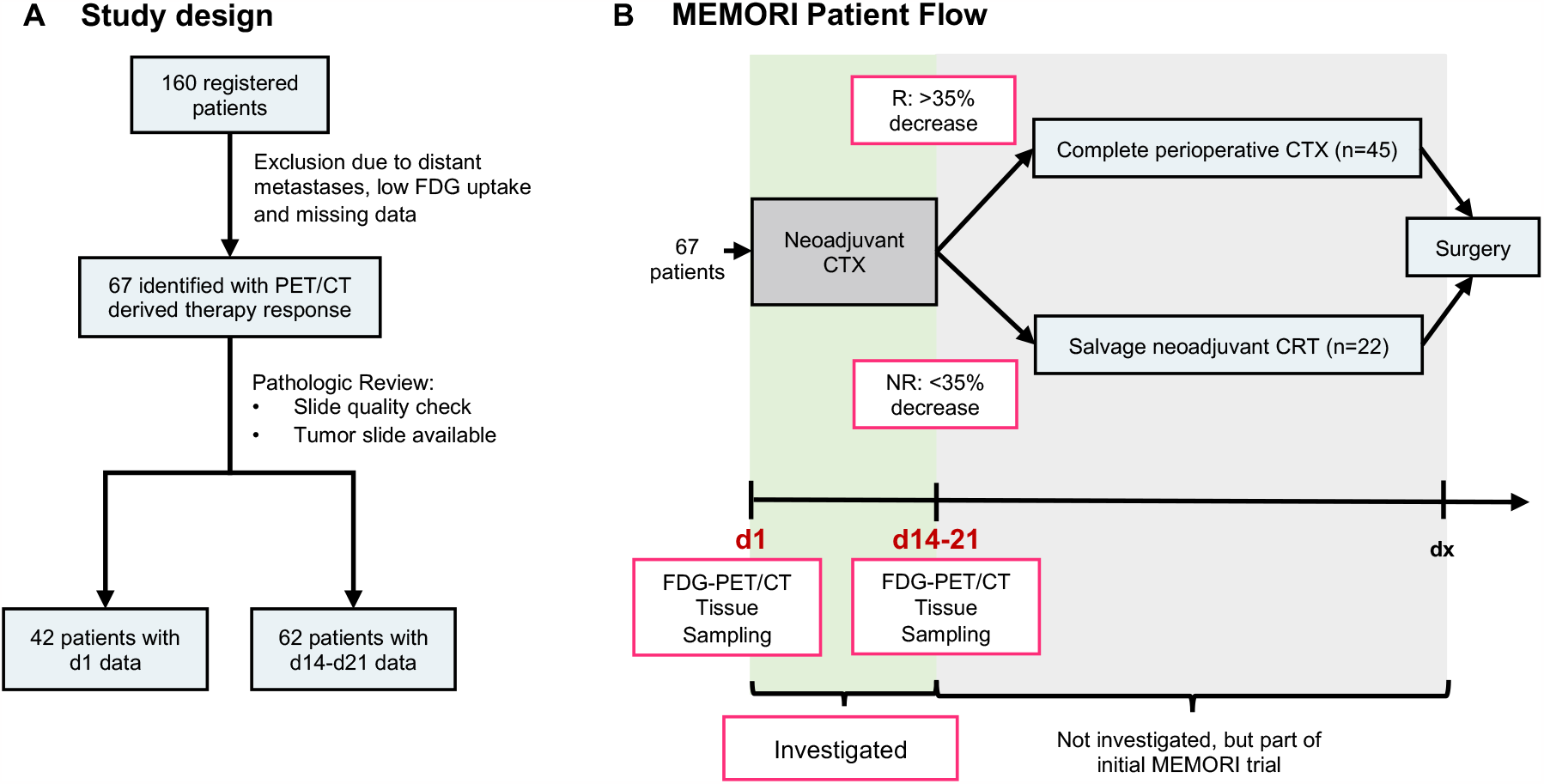
Study Design and Patient Flow. Study design for our retrospective recruited MEMORI cohort (A). The patients underwent neoadjuvant chemotherapy for 14-21 days, then being stratified in responder (R) and non-responder (NR) patients. We just consider the first cycle of CTX until patients had been stratified for our analysis (B). The complete MEMORI trial protocol also encompasses the analysis of therapy adaption based on FDG-PET/CT responder status in CTX or salvage chemoradiotherapy (CRT), which is not part of this investigation.

In total, 67 (TUM = 49, LMU = 39, UME = 9) patients were identified to have 22 per protocol treated non-responders with up to 4 tumor WSI per biopsy sampling. A trained board-certified pathologist reviewed all slides to ensure sufficient slide quality and annotated tumor areas. Subject to pathological review, 42 patients (TUM = 25, LMU = 8, UME = 9) with 152 WSI were included in the d1 analysis and 62 patients (TUM = 47, LMU = 6, UME = 9) with 204 WSI in the d14-21 analysis. Clinical characteristics, including age, sex, BMI, tumor grading, and TNM staging are listed in the Supplement (see Table S2).

### Development of the Deep Learning Model

Our proposed DL pipeline has three steps, as shown in Figure 2. The first step is to preprocess each scanned tissue specimen (WSI) to generate non-overlapping, quadratic patches of size 256 px and 4096 px, respectively, at 20 × magnification. For patch generation, tissue detection (Otsu^33^) is first performed, followed by Macenko stain normalization^15,34^. The second step is to extract histological image features out of the patches by using a deep learning based encoder network. Finally, all image features are aggregated by a decoder network into a patient feature vector for final treatment response prediction. Multiple neural networks for the encoder and decoder were combined to find the best possible prediction model. Rather than modeling temporal progression, the networks consider only WSI from a single sampling time (d1 vs. d14-21).

**Figure 2.**
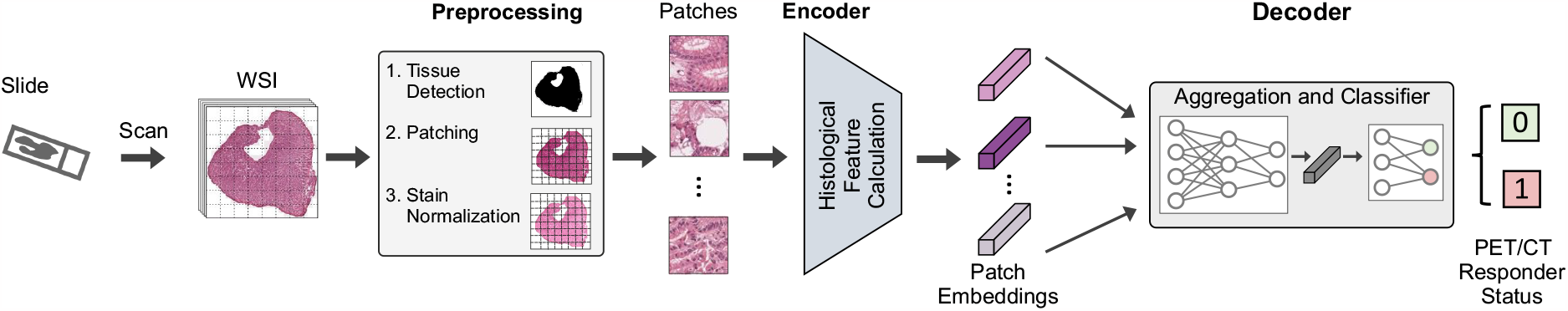
Deep learning pipeline for digitized tissue specimens to predict the treatment response determined by PET/CT. The WSI are first preprocessed to generate patches and then provided to the network. For each patch, image features are calculated, which are subsequently aggregated to predict the treatment response.

To extract image features from the patches, we examined two different state-of-the-art feature extraction methods: Convolutional neural networks (CNNs) and Vision Transformers^35^ (ViTs). Specifically, we used a modified ResNet50^36,37^ network as a CNN encoder network. Although high performance for histopathologic feature extraction was already achieved using ImageNet-based network weights^37,38^, we trained an additional ResNet50 encoder by utilizing the self-supervised representation learning algorithm SimTriplet^39^ on histopathological images to yield domain-specific features (e.g., morphological tissue features, cell features). As an alternative encoder structure, we used two recently published ViT architectures designed explicitly for histopathological feature extraction, called ViT-256 and ViT-4096, trained on 33 different cancer cases from The Cancer Genome Atlas (TCGA).^40,41^ The ViT-2ti6 network computes features of local cell clusters, that are combined by ViT-4096 into organizational units describing a tissue microenvironment.^40^

To aggregate the patch-wise feature vectors, we used two weakly-supervised approaches compromising multiple instance learning (MIL)^37,42^ and graph neural networks (GNNs)^38,43,44^. In the case of MIL, all patch features of one patient are aggregated by permutation invariant pooling operations, such as simple baseline mean and max pooling. To cover flexible patch contribution, we also used the clustering-constrained attention MIL (CLAM)^37^ network based on an interpretable attention mechanism. For each patch, an adaptive weight score is calculated depending on the patch information and the comparison to all remaining patches of a WSI. The score takes the relative patch importance for the models’ prediction into account. By using attention mechanisms in this way, we hope to improve the accuracy and interpretability of our results. In particular, various visualization techniques can be used to highlight the areas of the WSI that are most important for predicting therapy response. The authors of CLAM published weights for a network that was trained on TCGA data for non-small-cell-lung-cancer (NSCLC) subtyping, which we used for network initialization.

One drawback of the attention mechanism is the inability to deal with spatial context, due to the permutation invariant pooling operator. To overcome this limitation, we also tested a graph-based neural network, specifically Patch Graph Convolutional Network (PatchGCN)^38^. For this purpose, adjacent patch-wise feature vectors are connected via edges to build up a neighborhood graph. The edge weights between the individual feature vectors (modeling graph nodes) are again calculated via the attention mechanism. The feature vectors are successively aggregated by a neural network to a single patient vector, considering spatial patterns. In total, we used the following networks architectures:

- *Encoder:* ResNet50 (ImageNet), ResNet50 (SimSiam), ViT-256, ViT-4096
- *Decoder:* Max (MIL), Mean (MIL), CLAM (MIL), PatchGCN (GNN)

Further architectural details and training setups are given in the supplementary method section S1.

### Model Selection and Performance Evaluation

Among all encoder and decoder networks, we first needed to find the best combination for patient classification. We performed stratified Monte Carlo cross validation (MCCV, 75% train, 25% test set) on the patient level for each combination with 20 non-overlapping folds to estimate the models’ performance.^45^

As our models were designed to include just one timepoint, we performed model selection separately on d1 (pre-therapy) and d14-21 (on-therapy) samples. Given the best classification model structures retrieved by MCCV, models got trained and tested on data from TUM hospital and internally validated on data from UME and LMU hospitals. Besides internal validation, cross-timepoint evaluation was carried out on each final pre- and on-therapy classification model to examine if detected morphological features for treatment response are consistent.

Performance was assessed using four common metrics: the area under the receiver operating characteristic curve (AUROC), the area under the precision-recall curve retrieved by average precision (AUPRC), the balanced accuracy (B. Acc.) and Matthews correlation coefficient (MCC)^46^.

Qualitative heatmap visualizations based on the attention scores were created for attention-based networks. We selected the 250 highest attending patches of each patient in our validation set, clustered them with Density-Based Spatial Clustering (DBSCAN)^47^ (see supplementary Methods S2), and calculated two-dimensional representations using Uniform Manifold Approximation and Projection for Dimension Reduction (UMAP)^48^. The resulting clusters were analyzed by a trained board-certified pathologist to identify and interpret predictive regions and structures.

### Model Training

The ImageNet ResNet50 was used with default weights. In addition, we trained the ResNet50 with patches from the MEMORI dataset (d1 and d14-21) by using SimTriplet^39^. The pre-trained pan-cancer ViT-256 and ViT-4096 networks were not finetuned, as they already have been trained on 104 million cell-level images (ViT-256) and 408,218 tissue microenvironments (ViT-4096), suggesting sufficient feature extraction capabilities for histological images.

All decoder networks were optimized using Adam optimizer (Methods S2 in the Supplement). Training of the aggregation networks was performed at the WSI level with the patient label. The evaluation was always performed at the patient level including all available WSI.

For model selection with MCCV, we retrieved a dataset of 42 (27 R, 15 NR) patients with a total of 152 WSIs (105 R, 47 NR) for d1 and 62 (42 R, 20 NR) patients with a total of 204 WSIs (145 R, 59 NR) for d14-21. For internal d1 validation, 25 patients (17 R, 8 NR) with 90 WSI (67 R, 23 NR) were available from the TUM hospital for training and testing and a total of 17 patients (10 R, 7 NR) with 62 WSI (38 R, 24 NR) were available from the UME and LMU for final validation. For the internal d14-21 validation, the distribution is as follows: TUM 47 training and testing patients (32 R, 15 NR) with 147 WSI (106 R, 41 NR), UME and LMU 15 validation patients (10 R, 5 NR) with 47 WSI (29 R, 18 NR).

### Statistical Analysis

Data were analyzed between January and October 2022. Bootstrap resampling with 1000 repetitions was used to determine the 95% CIs of the test results. Statistical significance of clinical parameters was determined by Spearman’s rank correlation and T-test, both 2-sided, with a *P* < .05 significance level. Statistical analysis was carried out in Python (3.9.7) with SciPy^49^ (1.7.1), Pandas^50^ (1.3.4), and scikit-learn^51^ (1.1.2).

## Results

### Clinical Characteristics

The clinical parameters of the patients are presented in Table S2 and S3 in the supplementary material. We found no significant correlation between clinical parameters and treatment response to neoadjuvant CTX.

### Prediction Performance for Model Selection with MCCV

The MCCV classification results are summarized in Table 1. For pre-therapy (d1) experiments, both mean and max baseline pooling operators obtained random behavior (Table S4 in the Supplement), except for mean pooling in combination with the ViT-4096 encoder network with a mean AUROC of 0.72 (SD, 0.12). The three best-performing model combinations based on mean AUROC are the pre-trained CLAM network in combination with ResNet50 ImageNet (0.80, SD 0.14) and our ResNet50 SimTriplet encoder network (0.75, SD 0.15), respectively, and the graph-based PatchGCN network again with the ResNet50 SimTriplet encoder (0.74, SD 0.18).

**Table 1.** MCCV Results for model selection. Result format: Mean (SD). Abbreviations: AUROC, area under the receiver operating characteristic curve; AUPRC, area under the precision-recall curve; B. Acc., balanced accuracy; MCC, Matthews correlation coefficient; CLAM pre-trained, CLAM network pre-trained on TCGA NSCLC and finetuned on MEMORI data.

| Decoder | Encoder | AUROC | AUPRC | B. Acc. | MCC |
| --- | --- | --- | --- | --- | --- |
| **Pre-Therapy (d1) MCCV Results** |  |  |  |  |  |
| Mean | ViT-4096 | 0.72 (0.12) | 0.67 (0.17) | 0.55 (0.10) | 0.00 (0.00) |
| CLAM<br>Pre-trained | ResNet50 ImageNet | 0.80 (0.14) | 0.76 (0.14) | 0.72 (0.09) | 0.44 (0.18) |
|  | ResNet50 SimTriplet | 0.75 (0.15) | 0.69 (0.16) | 0.64 (0.15) | 0.28 (0.30) |
| PatchGCN | ResNet50 ImageNet | 0.72 (0.20) | 0.66 (0.18) | 0.50 (0.00) | 0.00 (0.00) |
|  | ResNet50 SimTriplet | 0.74 (0.18) | 0.68 (0.16) | 0.50 (0.00) | 0.00 (0.00) |
|  | ViT-256 | 0.58 (0.20) | 0.58 (0.19) | 0.56 (0.09) | 0.14 (0.21) |
| **On-Therapy (d14-d21) MCCV Results** |  |  |  |  |  |
| Max | ResNet50 ImageNet | 0.49 (0.09) | 0.35 (0.06) | 0.50 (0.00) | 0.00 (0.00) |
|  | ResNet50 SimTriplet | 0.54 (0.17) | 0.41 (0.12) | 0.50 (0.00) | 0.00 (0.00) |
|  | ViT-256 | 0.59 (0.08) | 0.39 (0.08) | 0.56 (0.08) | 0.15 (0.19) |
|  | ViT-4096 | 0.74 (0.13) | 0.59 (0.16) | 0.64 (0.12) | 0.28 (0.27) |
| Mean | ResNet50 ImageNet | 0.69 (0.12) | 0.65 (0.12) | 0.63 (0.09) | 0.36 (0.21) |
|  | ResNet50 SimTriplet | 0.81 (0.09) | 0.73 (0.12) | 0.69 (0.11) | 0.43 (0.21) |
|  | ViT-256 | 0.82 (0.09) | 0.75 (0.12) | 0.70 (0.09) | 0.46 (0.25) |
|  | ViT-4096 | 0.89 (0.08) | 0.83 (0.12) | 0.79 (0.10) | 0.61 (0.17) |
| CLAM | ResNet50 ImageNet | 0.72 (0.12) | 0.68 (0.12) | 0.58 (0.07) | 0.19 (0.19) |
|  | ViT-256 | 0.80 (0.10) | 0.74 (0.12) | 0.71 (0.11) | 0.48 (0.21) |
| CLAM<br>Pre-trained | ResNet50 ImageNet | 0.73 (0.14) | 0.64 (0.16) | 0.63 (0.14) | 0.29 (0.32) |
|  | ResNet50 SimTriplet | 0.86 (0.07) | 0.76 (0.12) | 0.68 (0.13) | 0.40 (0.28) |
| PatchGCN | ResNet50 ImageNet | 0.70 (0.14) | 0.59 (0.19) | 0.51 (0.02) | 0.02 (0.08) |
|  | ResNet50 SimTriplet | 0.84 (0.08) | 0.76 (0.10) | 0.70 (0.13) | 0.46 (0.25) |
|  | ViT-256 | 0.82 (0.11) | 0.73 (0.14) | 0.70 (0.14) | 0.43 (0.27) |

Overall, on-therapy (d14-21) networks showed superior predictive performance compared to d1 networks. Especially, the ViT-4096 encoder network in combination with mean pooling outperformed any other network combination for on-therapy biopsy samples with a mean AUROC of 0.89 (SD, 0.08). The pre-trained CLAM network with our ResNet50 SimTriplet encoder also achieved good results with an AUROC of 0.86 (SD, 0.07). Compared to d1 MCCV results, PatchGCN with ResNet50 SimTriplet encoder achieved a performance increase of 13% to 0.84 AUROC (SD, 0.10). In particular, the balanced classification performance for this setup improved from random guessing to 0.70 (SD, 0.13). AUROC distributions for the best networks are illustrated in supplementary Figure S4.

### Prediction Performance on Validation Cohort

Based on the MCCV test results, we selected the following four networks for internal validation: ViT-4096 with mean pooling, pre-trained CLAM network with ResNet50 (ImageNet, SimTriplet), and PatchGCN with ResNet50 SimTriplet encoder. The validation results are presented in Table 2. As previously reported for MCCV, the pre-trained CLAM network with ImageNet ResNet50 was the best-performing network for predicting the treatment response of neoadjuvant CTX based on pre-therapy biopsy samples. The network achieved a validation AUROC of 0.81 (95% CI, 0.61-1.00), AUPRC of 0.82 (95% CI, 0.61-1.00), balanced accuracy of 0.78 (95% CI, 0.60-0.94), and an MCC of 0.55 (95% CI, 0.18-0.88) on our internal validation cohort. Likewise, the combination of the ViT-4096 encoder and mean pooling achieved the best validation performance on the on-therapy biopsy samples, with an AUROC of 0.84 (95% CI, 0.64-1.00), AUPRC of 0.82 (95% CI, 0.56-1.00), balanced accuracy of 0.80 (95% CI, 0.63-1.00), and MCC of 0.71 (95% CI, 0.38-1.00).

**Table 2.** Validation Results on internal UME and LMU patient cohort. Result format: Mean (SD). Abbreviations: AUROC, area under the receiver operating characteristic curve; AUPRC, area under the precision-recall curve; B. Acc., balanced accuracy; MCC, Matthews correlation coefficient; CLAM pre-trained, CLAM network pre-trained on TCGA NSCLC and finetuned on MEMORI data.

| Decoder | Encoder | AUROC | AUPRC | B. Acc. | MCC |
| --- | --- | --- | --- | --- | --- |
| **Pre-Therapy (d1) Validation Results on UME and LMU Hospital data (n=17)** |  |  |  |  |  |
| Mean | ViT-4096 | 0.70 (0.46-0.94) | 0.61 (0.39-0.93) | 0.69 (0.48-0.88) | 0.38 (-0.04-0.75) |
| CLAM<br>Pre-trained | ResNet50 ImageNet | 0.81 (0.61-1.00) | 0.82 (0.61-1.00) | 0.78 (0.60-0.94) | 0.55 (0.18-0.88) |
|  | ResNet50 SimTriplet | 0.54 (0.30-0.79) | 0.48 (0.28-0.81) | 0.50 (0.50-0.50) | 0.00 (0.00-0.00) |
| PatchGCN | ResNet50 SimTriplet | 0.49 (0.32-0.67) | 0.43 (0.25-0.70) | 0.50 (0.50-0.50) | 0.00 (0.00-0.00) |
| **On-Therapy (d14-d21) Validation Results on UME and LMU Hospital data (n=15)** |  |  |  |  |  |
| Mean | ViT-4096 | 0.84 (0.64-1.00) | 0.82 (0.56-1.00) | 0.80 (0.63-1.00) | 0.71 (0.38-1.00) |
| CLAM<br>Pre-trained | ResNet50 ImageNet | 0.76 (0.50-1.00) | 0.67 (0.37-1.00) | 0.60 (0.50-0.80) | 0.38 (0.00-0.71) |
|  | ResNet50 SimTriplet | 0.74 (0.43-1.00) | 0.69 (0.33-1.00) | 0.55 (0.40-0.75) | 0.14 (-0.26-0.56) |
| PatchGCN | ResNet50 SimTriplet | 0.80 (0.56-1.00) | 0.80 (0.52-1.00) | 0.50 (0.50-0.50) | 0.00 (0.00-0.00) |

### Qualitative Performance on Validation Cohort

To interpret the DL model performance, we created representative attention heatmaps (Figure 3) for the CLAM network with ResNet50 ImageNet encoder for internal validation. For some patients, the attention heatmaps correlate strongly with the tumor regions. For others, high scores have also been assigned to peripheral tumor areas and surrounding tissue. Comparing the d1 heatmaps with the d14-21 heatmaps reveals that for d1 there is a more robust delineation between tumor regions and normal tissue. To further analyze the performance, we illustrated UMAP embeddings of the highest-attending patches along with DBSCAN clusters (Figure 3). For each cluster, representative tissue patches are shown in addition to exemplar low-attending patches. Regardless of the sample time, patches with low attention scores are mainly without important tissue information, e.g., corrupted/blurred patches, blood, detritus, and cell artifacts. Identified clusters in the d1 samples are one cluster with healthy squamous epithelium, two clusters with tumor, and one cluster consisting of macrophages and single tumor cells. In the d14-21 samples, we were again able to identify clusters of squamous epithelium and tumor areas. However, the clusters are not as well differentiated as in the d1 samples (e.g., cluster 3), and one cluster contains dysplastic columnar tissue. Additional visualizations using the ViT-256 and ViT-4096 encoders are provided in Figures S5 and S6 (Supplement).

**Figure 3.**
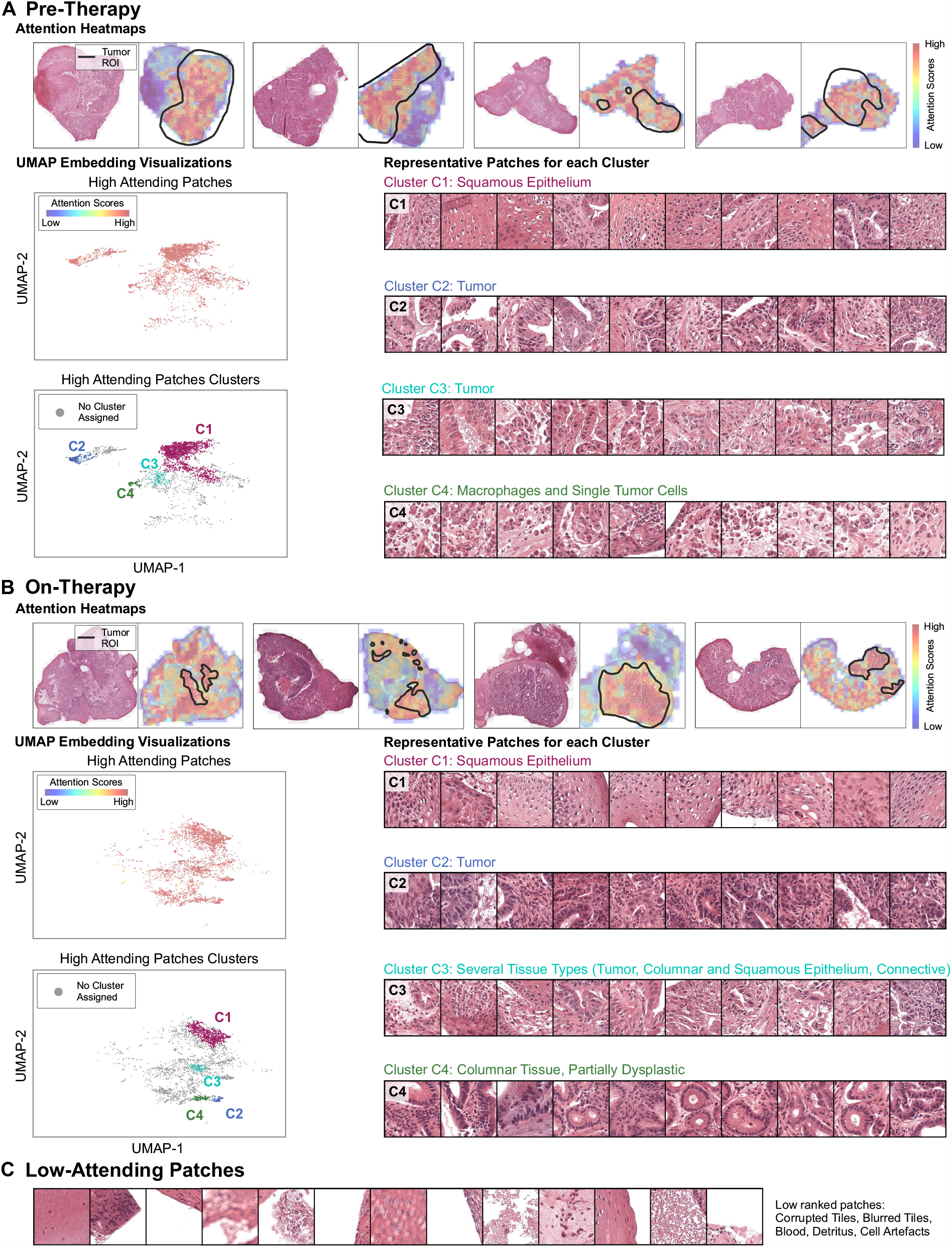
Attention-Score analysis using heatmaps and clustering for best-performing validation networks. The best network combination shown here is the pre-trained CLAM in combination with ResNet50 ImageNet, each for pre-therapy (A) and on-therapy (B) data. Exemplary attention heatmaps using relative attention scores are given along with tumor region of interest, drawn by a board-certified pathologist. The attention scores indicate the relative predictive importance of the tissue area. Highest attending patches (red heatmap regions) were used to generate cluster plots using the U-Map embedding algorithm. On the right, representative patches for identified clusters of high-attending patches are shown. Low-Attending patches (C) with no predictive importance are similar for both intervention times (pre-therapy vs. on-therapy).

### Prediction Performance on Cross-Timepoint Data

Testing the best-performing models from our internal validation sets on cross-timepoint data, we obtained a random AUROC classification performance of 0.48 (95% CI, 0.34-0.62) using d14-21 therapy biopsy samples on the best d1 model and 0.51 (95% CI, 0.35-0.65) vice versa.

## Discussion

This diagnostic study based on the prospective MEMORI trial is the first to evaluate the treatment response of GEJ cancer patients with histological slides using DL. We were able to select and train two networks achieving high accuracy on an internal validation cohort while maintaining high AUPRC values. These results suggest that H&E biopsy samples contain tissue morphologies indicating treatment response. The high accuracy with pre-therapy biopsy samples is of particular interest, which may support early patient stratification for therapy adjustment and justifies further tissue analysis in collaboration with pathologists. So far, pathologists have not been able to predict treatment response based on biopsy specimens alone. Also, besides PET/CT-based diagnostics, no known clinical parameters (e.g., TNM staging, tumor grading) show a significant correlation with treatment response.

Our work demonstrates that pre-trained models on WSI achieve superior performance. On the one hand, CLAM network pre-trained on TCGA NSCLC WSI achieves the best performance on the pre-therapy tissue samples and yields good results for the therapy tissue samples. The untreated GEJ tumor samples have morphologically similar structures to NSCLC tissue samples; Thus, the network generalizes. On the other hand, the combination of the pan-cancer TCGA pre-trained ViT-4096 encoder and mean-pooling achieves the best results for the on-therapy evaluation. Due to CTX, the treated samples have highly altered tissue, whereby pan-cancer pre-training generates more predictive features (see Figure S5 and S6 in the Supplement).

The attention heatmaps in Figure 3 aim to refuse shortcut learning and spurious attractors (e.g., staining differences) and to reveal key regions of the model. Both d1 and d14-d21 prediction models facilitate tumor regions and the surrounding tissue (tumor microenvironment). The clusters (Figure 3) confirm the visual heatmaps interpretation that mainly tumor areas are responsible for treatment response, but also healthy squamous epithelium has an influence. Low-attending patches, conversely, are patches without any meaningful tissue. The partially clear demarcation of the clusters in the pre-therapy specimens is remarkable, although esophageal adenocarcinoma is a very heterogeneous tumor, in which variability in the clusters would have been quite expected.

The inferior performance of the baseline models, together with the CLAM attention heatmaps and clusters (Figure 3), indicate that not a single small tissue section or the whole sample are equally important for prediction, but a complex interaction of the tumor with its microenvironment and the surrounding squamous epithelium is relevant.

In evaluating the best networks on image data from the other time, classification performance dropped dramatically. These performance drops are pathologically plausible since CTX does not selectively alter the tumor tissue. Instead, the tumor and surrounding tissue heterogeneity increase due to therapy, explaining the different patterns found. Thus, we showed that computationally accessible tissue changes result from the treatment.

The implications of our results are wide-ranging. Being able to detect non-responder patients based on pre-therapy WSI could significantly improve patient care and apply targeted therapy at an early therapy course. Nonetheless, we have only shown that patient stratification on digitized tissue samples is possible. The histopathological patterns that have led to this classification still need to be revealed and further research is crucial. In future work, it could be interesting to incorporate multiple imaging modalities (H&E, PET/CT) combined with blood and genetic testing in one multimodal model^52^ to test if prediction accuracy can be further enhanced to build one encompassing pre-screening test.

### Limitations

There are several limitations in this study. First, we just performed internal validation on a dataset acquired from two different German hospitals from patients of the MEMORI trial. In addition, our patient cohort is reasonably small, needing more extensive follow-up cohorts for external validation. All validation results must therefore be interpreted with caution. Second, our training label is based on the PET/CT SUVmax cut-off value. Thus, because no other method of determining treatment response is known, no conclusive comparison between the predictive performance of H&E biopsy images with our networks and PET/CT-guided treatment decisions^53^ is possible. Third, the qualitative interpretability of our approach is limited. Although we were able to show that our network does not perform shortcut learning, we can only interpret the learned features using our clustering. We were not able to identify new interpretable biomarkers. Fourth, we are currently limited to either using d1 or d14-21 images, not incorporating all available information in one network. Multimodal networks integrating different sources of information (H&E, PET/CT, blood test, genomic) may provide improved classification results with increased interpretability.

## Conclusions

In this diagnostic study, we developed two DL models to predict PET/CT treatment response status with high-resolution H&E biopsy sample images and achieved high accuracy even on the pre-therapy data on our internal validation set. This classifier could help with patient stratification for treatment adjustment at an early stage, if validated in prospective studies. We believe that this work provides an essential foundation to establish a new histological diagnostic system.

## Supporting information

Tripod Statement

## Data Availability

The participants of this study did not give written consent for their data to be shared publicly. There is no additional data available.

## Data Sharing

The participants of this study did not give wriden consent for their data to be shared publicly. There is no additional data available.

