## Supplement for "Histology-based Prediction of Therapy Response to Neoadjuvant Chemotherapy for Esophageal and Esophagogastric Junction Adenocarcinomas Using Deep Learning"

**Supplementary Material**

**Methods S1.** Deep Learning Model Framework

**Methods S2.** Implementation Details and Configuration

**References.**

**Figure S1.** Machine Learning Pipeline with Encoder and Decoder Network

**Figure S2.** SimTriplet Self-Supervised Learning of the Encoder $f_{enc}$

**Figure S3.** Boxplots of MCCV AUROC Results for the 4 Best-Performing Encoder-Decoder Combinations

**Figure S4.** ViT-256 Attention Heatmaps for Selected Attention Heads ($Head_2$, $Head_3$)

**Figure S5.** ViT-4096 Attention Heatmaps for Selected Attention heads ($Head_1$, $Head_2$, $Head_3$)

**Figure S6.** Embedding Visualizations for CLAM network.

**Table S1.** Inclusion and Exclusion Criteria of the MEMORI study

**Table S2.** Clinical Parameters of the MEMORI Study Cohort

**Table S3.** Clinical Parameters of the MEMORI Study Cohort, Split by Hospital

**Table S4.** Complete Pre-Therapy MCCV Results for Model Selection

**This supplementary material has been provided by the authors to give readers additional information about their work.**

### Methods S1. Deep Learning Model Framework

**1. Preliminaries**
For the sake of simplicity, the deep learning model framework is described for the case that one patient is just having one whole slide image. Let

$$\boldsymbol{w}^q \in R^{H\times W\times 3}, q \in [1, \ldots, N] \quad (1)$$

be a WSI $q$ with $(H, W)$ spatial dimensions and 3 color channels and $N$ be the number of all WSIs. Then the set off all WSI is denoted with $\mathcal{W} = \{\boldsymbol{w}^q, \ldots, \boldsymbol{w}^N\}$ . During Preprocessing, each WSI is divided into a set of non-overlapping quadratic patches

$$\mathcal{P}^q = \{\boldsymbol{p}_k^q\}, \boldsymbol{p}_k^q \in R^{S\times S\times 3}, k \in [1, \ldots, K^q], \quad (2)$$

with $K^q$ denoting the number of patches for the WSI $q$. Differing from equation (2), the extracted patches can also be denoted by

$$\mathcal{P}^q = \{\boldsymbol{p}_{i,j}^q\}, \boldsymbol{p}_{i,j}^q \in R^{S\times S\times 3}, \quad (3)$$

with $i$ and $j$ the row and column position of the patch in a two-dimensional grid to emphasize the spatial relation between patches. In our approach, the encoder $f_{\text{enc}}$ first maps a patch $\boldsymbol{p}_k^q$ into a feature vector $\boldsymbol{h}_k^q$ with

$$\boldsymbol{h}_k^q = f_{\text{enc}}(\boldsymbol{p}_k^q), \boldsymbol{h}_k^q \in R^{n_{\text{emdb}}}. \quad (4)$$

All $K^q$ patch embeddings of one WSI $q$ define the set WSI $\mathcal{H}^q = \{\boldsymbol{h}_1^q, \ldots, \boldsymbol{h}_{K^q}^q\}$. The aggregation function $f_{\text{agg}}$ then maps the set $\mathcal{H}^q$ to the corresponding slide label

$$\widehat{y^q} = f_{\text{agg}}(\mathcal{H}^q), \widehat{y^q} \in 0,1, \quad (5)$$

which, in fact, is the predicted PET/CT treatment outcome of the patient. The core of our framework consists of the functions $f_{\text{enc}}$ (eq. 4) and $f_{\text{agg}}$ (eq. 5). Since both functions are parametrized by neural networks, training them together in an end-to-end setting would theoretically be possible. Practically, several challenges occur: First, the amount of training data is limited. Typically, the number of slides for a study just includes several hundreds of slides with available training endpoints. Second, the number $K^q$ of patches in one WSI is too high (up to several thousand patches) for parallel processing in a single batch on current GPU hardware. Therefore, we stake to the solution of using dedicated encoders $f_{\text{enc}}$ that are not trained in conjunction with the aggregation networks $f_{\text{agg}}$. The advantage is that the embedding vectors $\boldsymbol{h}_k^q$ can be precalculated before training $f_{\text{agg}}$. Since the dimension $n^{\text{embd}}$ is significantly smaller than the original patch shape $R^{S\times S\times 3}$, the aggregation networks can be trained on all embedding vectors of one patient in parallel.

**2. Encoder: Networks for Feature Extraction**
In this work, we used two types of encoder networks: ResNet50[1] as a convolutional neural network based encoder network and two types of Vision Transformers[2] (ViT-256, and ViT-4096) published by Chen et al.[3]. Both encoder types are illustrated in eFigure 2.

**ResNet50:** Besides the ImageNet pretrained ResNet50 network, we used SimTriplet[4] as self-supervised contrastive learning to train a domain-specific ResNet50 network. An overview of the SimTriplet approach is given in eFigure 3. The SimTriplet architecture takes as input three image patches: Two views of a central patch and one view of an adjacent patch. During the forward pass, all views are processed with the same shared encoder $f_{\text{enc}}$ (ResNet50 without classification head) and a multi-layer FFN projection head, followed by a prediction head $f_{\text{pred}}$. The core assumption is that all three augmented views are positive views, meaning that they describe the same tissue phenotype and the similarity between their embedding vectors should be maximized. Cosine similarity $\mathcal{D}$ is used as the similarity metric. The total loss is assembled by a weighted sum (weight factor $\alpha = 0.7$) incorporating the intra-patch similarity and the inter-patch similarity:

$$\begin{aligned}
\mathcal{L} &= \alpha\mathcal{L}_{\text{intrasample}} + (1-\alpha)\mathcal{L}_{\text{intersample}} \\
\mathcal{L}_{\text{intrasample}} &= \frac{1}{2}\mathcal{D}(h_1', h_2) + \frac{1}{2}\mathcal{D}(h_2', h_1) \\
\mathcal{L}_{\text{intersample}} &= \frac{1}{2}\mathcal{D}(\boldsymbol{h}_2', \boldsymbol{h}_3) + \frac{1}{2}\mathcal{D}(\boldsymbol{h}_3', \boldsymbol{h}_2) \\
\mathcal{D}(\boldsymbol{x}, \boldsymbol{y}) &= -\frac{\boldsymbol{x}}{\|\boldsymbol{x}\|_2} \cdot \frac{\boldsymbol{y}}{\|\boldsymbol{y}\|_2}
\end{aligned} \quad (6)$$

To make use of already pretrained aggregation networks, we used a modified ResNet50 implementation[5] with additional spatial pooling to get an embedding vector with $1024$ features for each quadratic $256$ px patch.

**ViT256 and ViT-4096:** ViTs are token-based networks that decompose an input image into visual tokens and map them into an embedding vector using a Transformer network, where the spatial interaction of the visual tokens is captured by positional encoding.[2] Given a default token size of $16$ px $\times$ $16$ px and a resolution of $256$ px of an quadratic input patch acquired at $20\times$ magnification, the token size is approximately the size of a cell or other cell-like structures.[3] This property is exploited by the ViT-256 network to compute a feature vector from local cell clusters out of a $256$ px large image section. We follow the implementation of Chen et al.[3] for ViT-256, retrieving a $384$-dimensional feature vector for each patch. ViT-4096 is used to combine the local cell clusters of ViT-256 into organizational units that describe a tissue microenvironment with the size of $4096$ px $\times$ $4096$ px.[3] The Substantial advantage of ViT-4096 is that due to the histopathologic specified network design and the cell-level ViT-256 as first input network, large interconnected tissue regions can be aggregated by considering spatial interactions. Thus, a $4096$ px-sized input patch is compressed into a $192$-dimensional feature vector. Technically, the ViT-256 output class token of an input patch ($256$ px) is used as one input token for the ViT-4096 network.
The ViT-256 and ViT-4096 network were trained with the self-supervised training technique DINO[6] on $10{,}678$ WSI from $33$ different cancer types out of the cancer genome atlas (TCGA), resulting in $104$ M $256$ px $\times$ $256$ px training patches for ViT-256 and $408{,}218$ $4096$ px $\times$ $4096$ px training patches for ViT-4096.[3] Network weights are publicly available by the authors.

**3. Decoder: Patient-Level Aggregation**
We considered three different types of aggregation functions, shown in eFigure 2: Multiple instance learning based aggregation functions[5,7], and PatchGCN[8] as a graph-based approach. All approaches calculate a patient feature vector $\boldsymbol{z}^q$ followed by a simple linear layer classifier to derive the patient label $\widehat{y^q}$.

**Multiple Instance Learning:** The structure of the used MIL network $f_{\text{agg}}$ is related to the CLAM network[5]. The input feature set $\mathcal{H}^q$ can be represented as a matrix $\boldsymbol{H}^q = [\boldsymbol{h}_1^q, \dots, \boldsymbol{h}_{K^q}^q], \boldsymbol{H}^q \in R^{n_{\text{embd}} \times K^q}$. The embedding dimension is first downscaled by a linear layer, followed by the MIL pooling layer to retrieve the patient feature vector $\boldsymbol{z}^q$:

$$\boldsymbol{z}^q = f_{\text{pool}}(\boldsymbol{W}\boldsymbol{H}^q) = f_{\text{pool}}([\boldsymbol{z}_1^q, \dots, \boldsymbol{z}_{K^q}^q]) \quad (6)$$

The non-trainable maximum and mean-operator serve as baseline operators:

$$max: \forall_{m=1,\dots,n_{\text{down}}}: \boldsymbol{z}_m^q = \max_{k=1,\dots,K} \boldsymbol{z}_{km}^q \qquad mean: \boldsymbol{z}^q = \frac{1}{K^q}\sum_{k=1}^{K^q} \boldsymbol{z}_k^q \quad (7)$$

However, they have the disadvantage of fixed parametrization. To overcome this, Ilse et al.[7] proposed an interpretable MIL approach based on an attention mechanism:

$$\boldsymbol{z}^q = \sum_{k=1}^{K^q} a_k^q \boldsymbol{z}_k^q, \text{with } a_k^q = \frac{\exp\left\{W_{\text{attn}}^T\left(\tanh(V\boldsymbol{z}_k^q) \odot \sigma(U\boldsymbol{z}_k^q)\right)\right\}}{\sum_{j=1}^{K^q}\exp\left\{W_{\text{attn}}^T\left(\tanh(V\boldsymbol{z}_k^q) \odot \sigma(U\boldsymbol{z}_k^q)\right)\right\}} \quad (8)$$

The operator $\odot$ is the element-wise multiplication, $\tanh(\cdot)$ the hyperbolic tangent and $\sigma(\cdot)$ the sigmoid activation function. The operations in eq. (8) are widely known as attention mechanism and the weights $a_k^q$ are called attention scores.[7] Due to the adaptive weight calculation, different weights can be assigned to every patch within a bag. This helps in finding the key patches of a WSI. Since the attention scores always sum up to 1, they are independent of the bag size.[7]
Lu et al.[5] (CLAM) extended the attention mechanism by including an instance clustering over identified representative tissue patches to receive more discriminative feature embeddings. For every class $c$ of the classification task, one separate attention branch is used to calculate $c$ patient feature vectors. The final classifier then calculates a score for each class $c$ and applies a softmax activation function to predict the treatment response probability distribution.[5] The most and least attending patches of each class, determined by their attention scores $a_{k,c}^q$ are used for a sperate clustering constrain to maximize the decision boundary between all classes. The intuition is that patches with high attention scores are expected to be important for the slide label prediction, whereas patches with low attention scores are

expected to be contrary evidence for slide label.[5] The formulas are valid for a patient $q$ having one WSI $q$. Since a patient can have multiple WSI from different biopsy locations, in practice we incorporate all WSIs patches from one patient $q$ into one set $\mathcal{H}^q$. This is possible because $f_{\text{pool}}$ in the MIL setting is permutation invariant.

**PatchGCN:** We followed the implementation of Chen et al.[8]. The basis of this network structure is the formation of a graph $G^q$ at the WSI level. Each feature vector $\boldsymbol{h}_{i,j}^q \in \mathcal{H}^q$ is one node of $G^q$, such that each histology image patch corresponds to one graph node.[8] The node feature matrix is denoted with $\boldsymbol{H}^q = [\boldsymbol{h}_1^q, \dots, \boldsymbol{h}_{K^q}^q], \boldsymbol{H}^q \in R^{n_{\text{embd}} \times K^q}$ .The edges are connected between adjacent image patches from the true spatial coordinates of the WSI and the adjacency matrix $\boldsymbol{A}^q$ is built with a k-NN ($k = 8$) algorithm, comparable to a $3 \times 3$ receptive field of a CNN.[8] Thus, the graph is defined by $G^q = (\boldsymbol{H}^q, \boldsymbol{A}^q)$. The graph neural network is parametrized using a graph convolutional neural network (GCN) by stacking $L = 4$ graph convolutional layers. Each layer consists of a message-passing step where information between the nodes and their neighbours (connected by edges) are shared and a permutation invariant aggregation function (attention based) that aggregates all received messages of each node. After the last graph convolution layer, a global attention based pooling layer similar to Ilse et al.[7] (cf. eq. 8) is used to retrieve the classification vector $\boldsymbol{z}^q$. To extend this to patient with multiple WSIs, the graphs of one patient can be assembled to one big graph by extending the node feature matrix and adjacency matrix such that each WSI graph $G^q$ is an isolated subgraph.

### Methods S2. Implementation Details and Configuration

#### 1. Implementation Details

All code used for this work is implemented in Python. The neural network models are implemented using PyTorch[9] for Python. PyTorch is a Python framework that allows to define and train neural networks on CPU and GPU hardware. WSI files are handled by using the OpenSource framework OpenSlide[10]. For experiment tracking and visualizations, we used the local running solution Tensorboard to achieve data sovereignty. Annotations are performed using the OpenSource Software QuPath[11].

#### 2. Network Configurations and Hyperparameters

If not otherwise stated, default network configurations and training procedures as in the original publications have been used. For the ViT-256 and ViT-4096 backbone, we used the ViT-256-S and ViT-4096-XS pretrained networks with the following configurations:

- ViT-256 = (patch-size: $256$, token-size: $16$, $n_{\text{embd}}$: $384$, layers: $12$, heads: $6$, MLP-Size: $4 \cdot 384$)
- ViT-4096 = (input-embedding: $384$, $n_{\text{embd}}$: $192$, layers: $6$, heads: $6$, MLP-Size: $4 \cdot 192$)

For training the aggregation networks, we used Adam optimizer for $100$ epochs with early stopping after 25 epochs, weight decay and learning rate scheduling, with the following hyperparameters: $\eta = 0.005, \beta_1 = 0.85, \beta_2 = 0.9, \lambda = 0.0001, \eta \leftarrow 0.95\eta$. In addition, dropout ($p_{\text{dropout}} = 0.25$) is used.
Balanced accuracy and MCC were calculated by using an unoptimized classification threshold of 0.5.

#### 3. DBSCAN-Clustering

Clustering of high-attending patches was performed with the DBSCAN[12] algorithm. From each patient, the $250$ highest patches were selected to build the patch-clustering dataset. First, the high-dimensional patch features were reduced to a $50$-dimensional feature vector with principal component analysis (PCA), followed by UMAP[13] ($n_{neighbours} = 7$, $min_{dist} = 0.1$, $metric = Euclidean$) reduction to a two-dimensional embedding vector. Clustering was performed on the downscaled PCA embedding vectors across all selected patches with DBSCAN ($\epsilon = 0.3$, $min_{samples} = 40$, $metric = Euclidean$). Simultaneously, only clusters with patches from more than $4$ patients were taken into account, and clusters with less than $4$ patients were discarded.

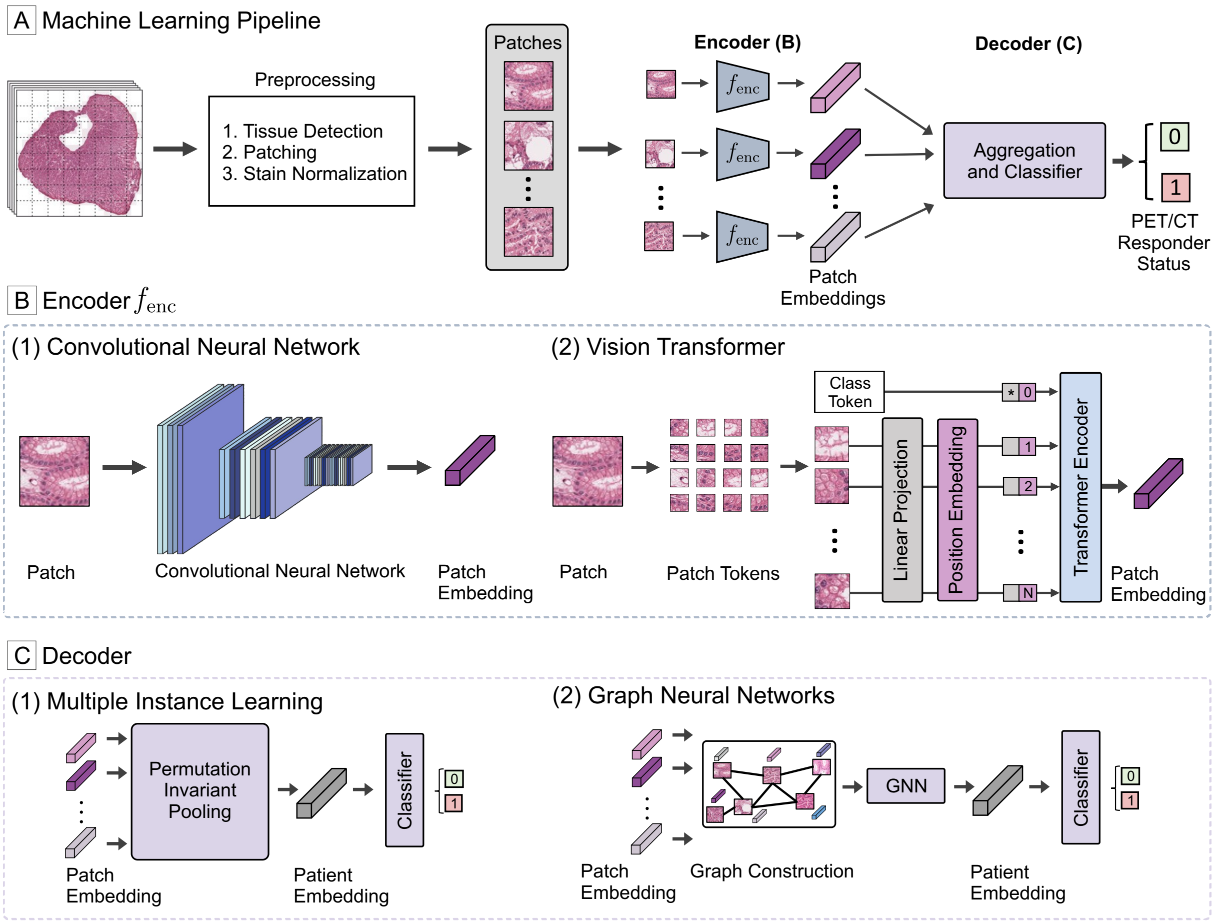


**Figure S1. Machine Learning Pipeline with Encoder and Decoder Network**
Deep learning pipeline to predict the PET/CT treatment response. A) Pipeline consisting of preprocessing, patch-encoding, and decoder network for aggregation. B) CNN and ViT-based encoder structures used for calculating patch features and C) MIL, graph-based, and ViT decoder network architectures.

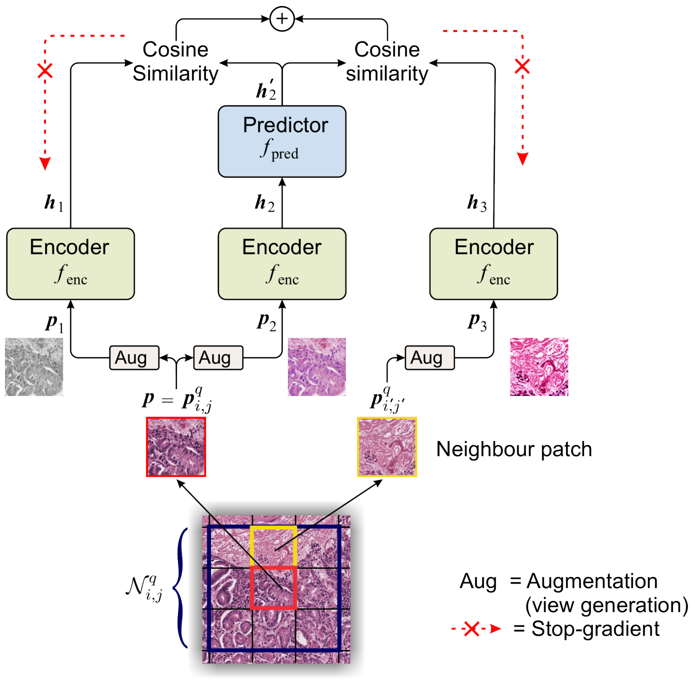


**Figure S2. SimTriplet[4] Self-Supervised Learning of the Encoder $f_{enc}$**

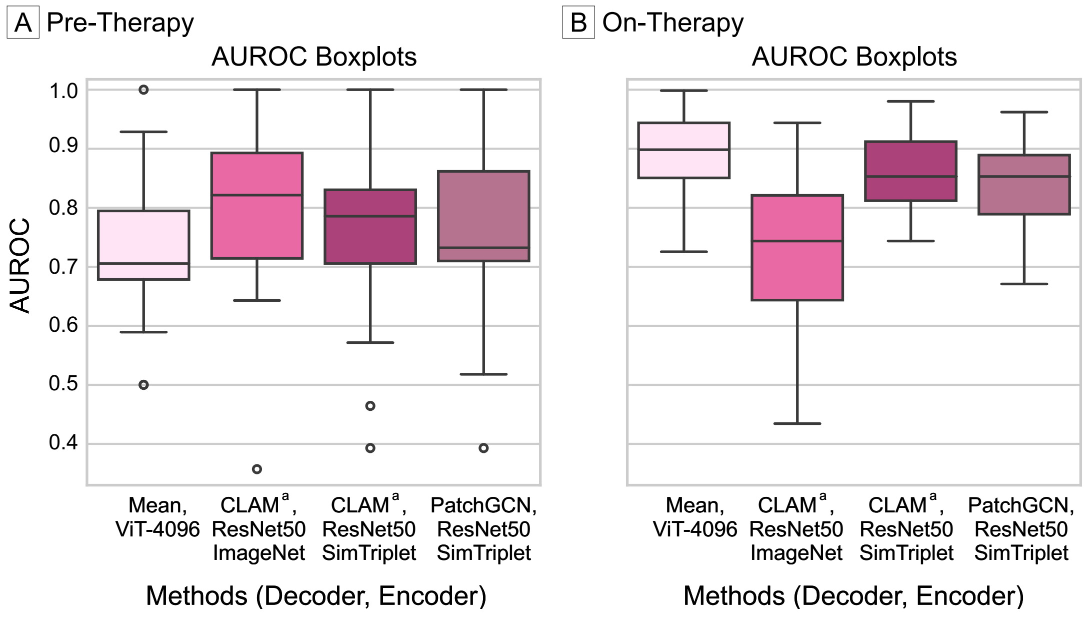


**Figure S3. Boxplots of MCCV AUROC Results for the 4 Best-Performing Encoder-Decoder Combinations**

Abbreviations: CLAM, CLAM network pretrained on TCGA NSCLC dataset; AUROC, area under the receiver operating characteristic curve. [a]Pretrained on TCGA.

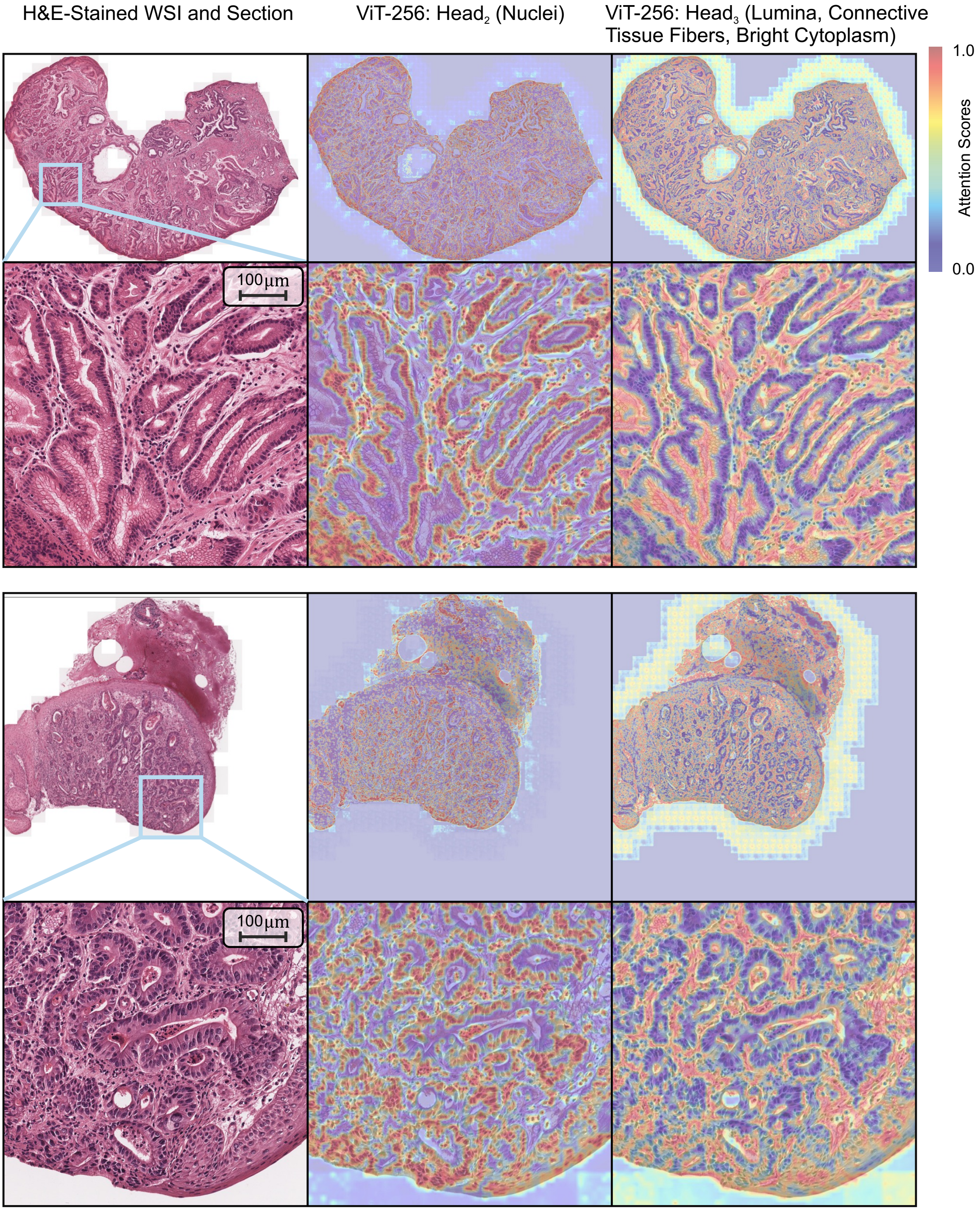


**Figure S4. ViT-256 Attention Heatmaps for Selected Attention Heads (Head$_2$, Head$_3$)**

The attention weights of Head$_2$ clearly mark cell nuclei, whereas Head$_3$ focuses on lumina, connective tissue fibers, and bright cytoplasm. Interestingly, Head$_3$ is able to distinguish between these tissue parts and empty background regions, thus taking cellular context into account and not just focusing on white/bright color.

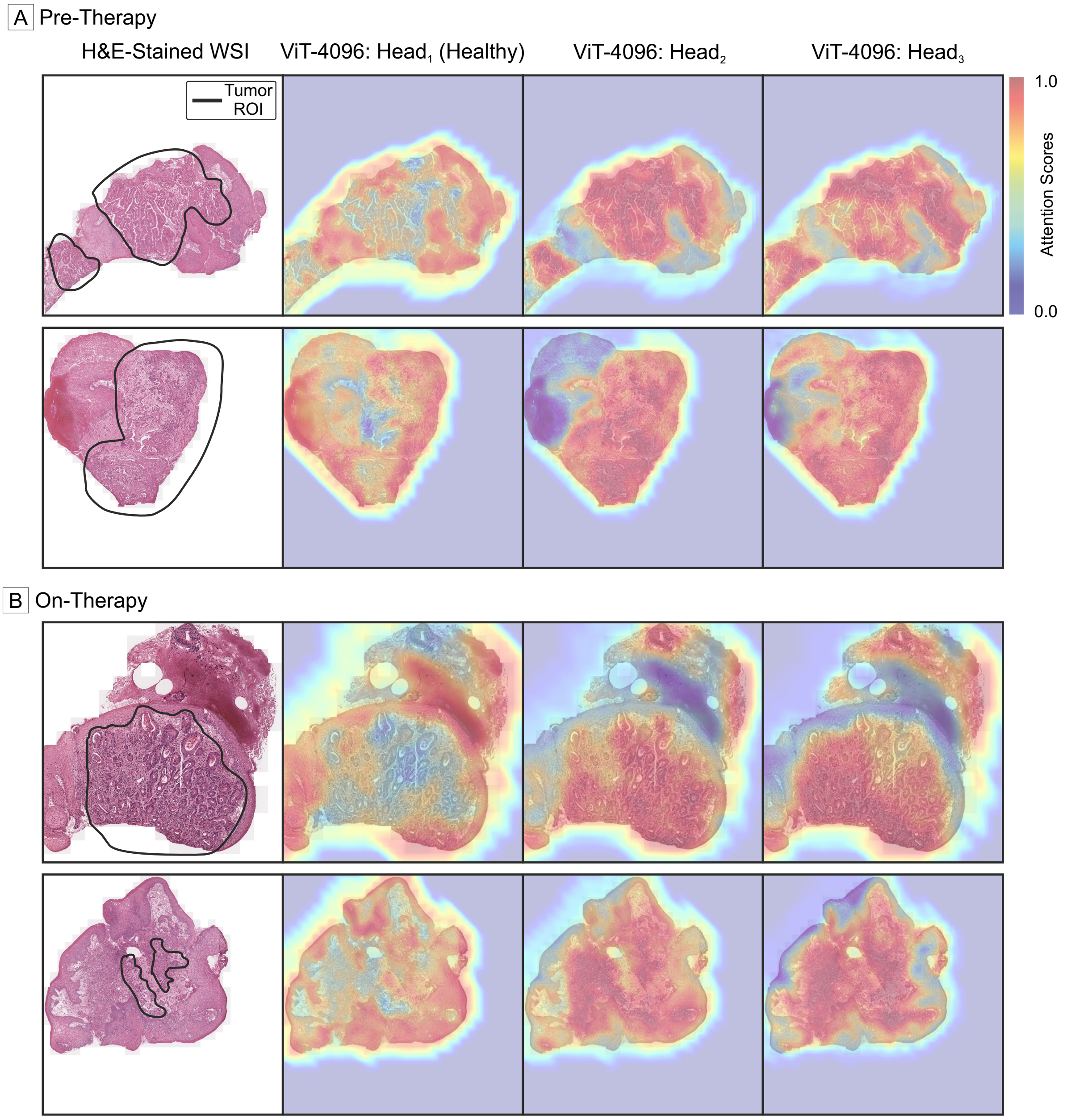


**Figure S5. ViT-4096 Attention Heatmaps for Selected Attention Heads ($Head_1$, $Head_2$, $Head_3$)**

The attention weights of $Head_1$ mostly localize healthy tissue and tissue without inflammatory cells. Both $Head_2$ and $Head_3$ assign high scores to the tumor regions with surrounding inflammatory tissue, with $Head_2$ primarily assessing the tumor regions. $Head_3$ includes more surrounding tissue. Especially for the on-therapy samples, the demarcations between tumor and healthy tissue are not clearly visible due to the tissue changes (reactive tissue, dysplasia) caused by neoadjuvant CTX. Nevertheless, the attention heatmaps reveal that the ViT-256 and ViT-4096 encoders calculate reasonable, pathologically interpretable tissue representations.

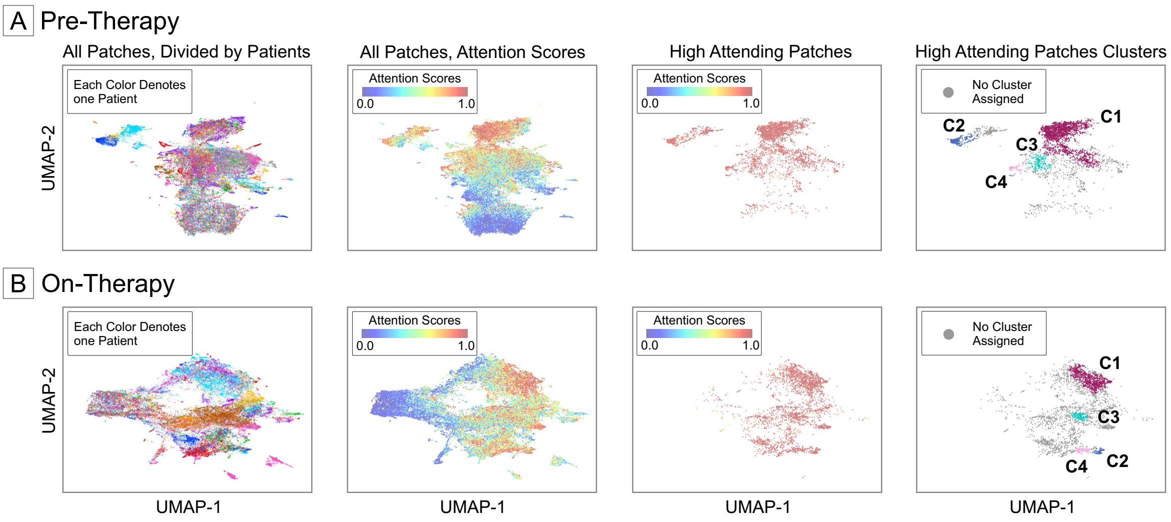


**Figure S6. UMAP Embedding Visualizations for CLAM network.**
Shown are UMAP representations of the patch embeddings with different color overlays. The used network combination for pre-therapy (A) and on-therapy (B) comparison is the TCGA pretrained CLAM network in combination with ResNet50 ImageNet.

### Table S1. Inclusion and Exclusion Criteria of the MEMORI Study

| Inclusion Criteria | Exclusion Criteria |
|---|---|
| • Histologically confirmed GEJ I-III<br>• Potentially R0-resectable GEJ and primary tumor category UT2-4<br>• Functional operability: Exclusion of OP-limiting comorbidities<br>• Intense FDG tracer uptake of the tumor during Baseline PET/CT examination and thus suitability for monitoring and early response prediction by FDG-PET ([ 18F]-FDG uptake in the tumor at baseline > 1.35 x liver SUV + 2 x standard deviation of the liver SUV)<br>• Performance status (ECOG) 0 or 1<br>• Age: ≥ 18<br>• Creatinine clearance > 60ml/min measured in a 24 h urine or calculated with the Cockgroft-Gault formula<br>• Bilirubin ≤ 1.5 times upper limit of normal, serum transaminases (GOT/GPT) ≤ 3 times ULN<br>• Leukocytes ≥ 3.5 g / l, platelet ≥ 100 g / l<br>• Negative pregnancy test (determination of beta-HCG in urine or serum) in women of childbearing potential<br>• A signed consent form after implementation of medical education | • Existing distant metastases (M1b)<br>• Tumor infiltration into the tracheobronchial system<br>• Previous radiotherapy targeted at the thorax<br>• Lack of ability of the patient to adhere to the protocol rules<br>• Manifest heart failure despite optimal medication> NYHA I<br>• Existing angina pectoris at rest or undergoing stress without clarification via interventional cardiology and/or myocardial infarction within last 6 months<br>• Existing pregnancy or lactation<br>• Childbearing or fertility without using recognized safe methods of contraception<br>• Coexisting other malignant diseases with the exception of a non-melanomatuous, localized skin tumor or carcinoma in situ of the cervix<br>• Absence of a signed consent form |

### Table S2. Clinical Parameters of the MEMORI Study Cohort

Abbreviations: R, Responder; NR, Non-Responder

| **Variables** | **Complete Dataset ($n = 67$)** | | | |
|---|---|---|---|---|
| | Total ($n = 67$) | R ($n = 45$) | NR ($n = 22$) | $P$ |
| Gender | | | | .56 |
| Female | 7 (0.10) | 4 (0.09) | 3 (0.14) | |
| Male | 60 (0.90) | 41 (0.91) | 19 (0.86) | |
| Age | 59.46 (9.68) | 59.73 (8.53) | 58.91 (11.89) | .75 |
| BMI | 27.33 (5.02) | 26.63 (5.38) | 28.72 (3.96) | .11 |
| Grading | | | | .59 |
| GX | 4 (0.06) | 1 (0.02) | 3 (0.14) | |
| G1 | 6 (0.09) | 4 (0.09) | 2 (0.09) | |
| G2 | 28 (0.42) | 21 (0.47) | 7 (0.32) | |
| G3 | 29 (0.43) | 19 (0.42) | 10 (0.45) | |
| cT | | | | .43 |
| TX | 2 (0.03) | 1 (0.02) | 1 (0.05) | |
| T2 | 7 (0.10) | 4 (0.09) | 3 (0.14) | |
| T3 | 55 (0.82) | 37 (0.82) | 18 (0.82) | |
| T4 | 3 (0.04) | 3 (0.07) | 0 (0.00) | |
| cN | | | | .78 |
| NX | 44 (0.66) | 31 (0.69) | 13 (0.59) | |
| N0 | 7 (0.10) | 4 (0.09) | 3 (0.14) | |
| N1 | 13 (0.19) | 9 (0.20) | 4 (0.18) | |
| N2 | 3 (0.04) | 1 (0.02) | 2 (0.09) | |
| cM | | | | .49 |
| M0 | 66 (0.99) | 44 (0.98) | 22 (1.00) | |
| M1 | 1 (0.01) | 1 (0.02) | 0 (0.00) | |
| Resection | | | | .92 |
| RX | 2 (0.03) | 0 (0.00) | 2 (0.09) | |
| R0 | 62 (0.93) | 43 (0.96) | 19 (0.86) | |
| R1 | 3 (0.04) | 2 (0.04) | 1 (0.05) | |
| ypT | | | | .72 |
| T0 | 4 (0.06) | 2 (0.04) | 2 (0.09) | |
| T1 | 13 (0.19) | 9 (0.20) | 4 (0.18) | |
| T2 | 18 (0.27) | 14 (0.31) | 4 (0.18) | |
| T3 | 32 (0.48) | 20 (0.44) | 12 (0.55) | |
| ypN | | | | .91 |
| NX | 1 (0.01) | 0 (0.00) | 1 (0.05) | |
| N0 | 38 (0.57) | 25 (0.56) | 13 (0.59) | |
| N1 | 19 (0.28) | 16 (0.36) | 3 (0.14) | |
| N2 | 5 (0.07) | 3 (0.07) | 2 (0.09) | |
| N3 | 4 (0.06) | 1 (0.02) | 3 (0.14) | |
| ypM | | | | .97 |
| MX | 17 (0.25) | 12 (0.27) | 5 (0.23) | |
| M0 | 50 (0.75) | 33 (0.73) | 17 (0.77) | |

### Table S3. Clinical Parameters of the MEMORI Study Cohort, Split by Hospital

Abbreviations: R, Responder; NR, Non-Responder; MRI, Klinikum rechts der Isar; LMU, Klinikum der Universität München; UKE, Universitätsklinikum Essen

| Variables | MRI ($n = 49$) | | | | LMU ($n = 9$) | | | | UKE ($n = 9$) | | | |
|---|---|---|---|---|---|---|---|---|---|---|---|---|
| | Total ($n = 49$) | R ($n = 34$) | NR ($n = 15$) | $P$ | Total ($n = 9$) | R ($n = 5$) | NR ($n = 4$) | $P$ | Total ($n = 9$) | R ($n = 6$) | NR ($n = 3$) | $P$ |
| Gender | | | | .64 | | | | .29 | | | | .52 |
| Female | 5 (0.10) | 3 (0.09) | 2 (0.13) | | 1 (0.11) | 0 (0.00) | 1 (0.25) | | 1 (0.11) | 1 (0.17) | 0 (0.00) | |
| Male | 44 (0.90) | 31 (0.91) | 13 (0.87) | | 8 (0.89) | 5 (0.00) | 3 (0.75) | | 8 (0.89) | 5 (0.83) | 3 (1.00) | |
| Age | 60.04 (10.2) | 60.12 (8.88) | 59.87 (13.09) | .94 | 60.67 (7.00) | 58.80 (8.17) | 63.00 (5.35) | .41 | 55.11 (8.58) | 58.33 (7.92) | 48.67 (6.66) | .11 |
| BMI | 27.63 (4.81) | 27.01 (4.81) | 28.99 (4.68) | .19 | 25.89 (3.87) | 23.73 (3.74) | 28.59 (1.95) | .05 | 27.18 (7.14) | 26.98 (8.95) | 27.58 (1.84) | .91 |
| Grading | | | | .55 | | | | .10 | | | | .14 |
| GX | 2 (0.04) | 1 (0.03) | 1 (0.07) | | 2 (0.22) | 0 (0.00) | 2 (0.50) | | 0 (0.00) | 0 (0.00) | 0 (0.00) | |
| G1 | 1 (0.02) | 1 (0.03) | 0 (0.00) | | 3 (0.33) | 1 (0.20) | 2 (0.50) | | 2 (0.22) | 2 (0.33) | 0 (0.00) | |
| G2 | 22 (0.45) | 16 (0.47) | 6 (0.40) | | 2 (0.22) | 2 (0.40) | 0 (0.00) | | 4 (0.44) | 3 (0.50) | 1 (0.33) | |
| G3 | 24 (0.49) | 16 (0.47) | 8 (0.53) | | 2 (0.22) | 2 (0.40) | 0 (0.00) | | 3 (0.33) | 1 (0.17) | 2 (0.67) | |
| cT | | | | .50 | | | | .53 | | | | .52 |
| TX | 0 (0.00) | 0 (0.00) | 0 (0.00) | | 2 (0.22) | 1 (0.20) | 1 (0.25) | | 0 (0.00) | 0 (0.00) | 0 (0.00) | |
| T2 | 1 (0.02) | 1 (0.03) | 0 (0.00) | | 5 (0.56) | 2 (0.40) | 3 (0.75) | | 1 (0.11) | 1 (0.17) | 0 (0.00) | |
| T3 | 45 (0.92) | 30 (0.88) | 15 (1.00) | | 2 (0.22) | 2 (0.40) | 0 (0.00) | | 8 (0.89) | 5 (0.83) | 3 (1.00) | |
| T4 | 3 (0.06) | 3 (0.09) | 0 (0.00) | | 0 (0.00) | 0 (0.00) | 0 (0.00) | | 0 (0.00) | 0 (0.00) | 0 (0.00) | |
| cN | | | | .85 | | | | - | | | | 1.00 |
| NX | 38 (0.78) | 27 (0.79) | 11 (0.73) | | 5 (0.56) | 4 (0.80) | 1 (0.25) | | 1 (0.11) | 0 (0.00) | 1 (0.33) | |
| N0 | 5 (0.10) | 3 (0.09) | 2 (0.13) | | 0 (0.00) | 0 (0.00) | 0 (0.00) | | 2 (0.22) | 1 (0.17) | 1 (0.33) | |
| N1 | 5 (0.10) | 4 (0.12) | 1 (0.07) | | 4 (0.44) | 1 (0.20) | 3 (0.75) | | 4 (0.44) | 4 (0.67) | 0 (0.00) | |
| N2 | 1 (0.02) | 0 (0.00) | 1 (0.07) | | 0 (0.00) | 0 (0.00) | 0 (0.00) | | 2 (0.22) | 1 (0.17) | 1 (0.33) | |
| cM | | | | .51 | | | | - | | | | - |
| M0 | 48 (0.98) | 33 (0.97) | 15 (1.00) | | 9 (1.00) | 5 (1.00) | 4 (1.00) | | 9 (1.00) | 6 (1.00) | 3 (1.00) | |
| M1 | 1 (0.02) | 1 (0.03) | 0 (0.00) | | 0 (0.00) | 0 (0.00) | 0 (0.00) | | 0 (0.00) | 0 (0.00) | 0 (0.00) | |
| Resection | | | | .38 | | | | - | | | | .17 |
| RX | 2 (0.04) | 0 (0.00) | 2 (0.13) | | 0 (0.00) | 0 (0.00) | 0 (0.00) | | 0 (0.00) | 0 (0.00) | 0 (0.00) | |
| R0 | 45 (0.92) | 32 (0.94) | 13 (0.87) | | 9 (1.00) | 5 (1.00) | 4 (1.00) | | 8 (0.89) | 6 (0.00) | 2 (0.67) | |
| R1 | 2 (0.04) | 2 (0.06) | 0 (0.00) | | 0 (0.00) | 0 (0.00) | 0 (0.00) | | 1 (0.11) | 0 (0.00) | 1 (0.33) | |
| ypT | | | | .37 | | | | .63 | | | | .80 |
| T0 | 4 (0.08) | 2 (0.06) | 2 (0.13) | | 0 (0.00) | 0 (0.00) | 0 (0.00) | | 0 (0.00) | 0 (0.00) | 0 (0.00) | |
| T1 | 7 (0.14) | 6 (0.18) | 1 (0.07) | | 3 (0.33) | 1 (0.20) | 2 (0.50) | | 3 (0.33) | 2 (0.33) | 1 (0.33) | |
| T2 | 12 (0.24) | 10 (0.29) | 2 (0.13) | | 4 (0.44) | 3 (0.60) | 1 (0.25) | | 2 (0.22) | 1 (0.17) | 1 (0.33) | |
| T3 | 26 (0.53) | 16 (0.47) | 10 (0.67) | | 2 (0.22) | 1 (0.20) | 1 (0.25) | | 4 (0.44) | 3 (0.50) | 1 (0.33) | |
| ypN | | | | .84 | | | | .12 | | | | .24 |
| NX | 0 (0.00) | 0 (0.00) | 0 (0.00) | | 1 (0.11) | 0 (0.00) | 1 (0.25) | | 0 (0.00) | 0 (0.00) | 0 (0.00) | |
| N0 | 29 (0.59) | 20 (0.59) | 9 (0.60) | | 5 (0.56) | 2 (0.40) | 3 (0.75) | | 4 (0.44) | 3 (0.50) | 1 (0.33) | |
| N1 | 13 (0.27) | 10 (0.29) | 3 (0.20) | | 3 (0.33) | 3 (0.60) | 0 (0.00) | | 3 (0.33) | 3 (0.50) | 0 (0.00) | |
| N2 | 4 (0.08) | 3 (0.09) | 1 (0.07) | | 0 (0.00) | 0 (0.00) | 0 (0.00) | | 1 (0.11) | 0 (0.00) | 1 (0.33) | |
| N3 | 3 (0.06) | 1 (0.03) | 2 (0.13) | | 0 (0.00) | 0 (0.00) | 0 (0.00) | | 1 (0.11) | 0 (0.00) | 1 (0.33) | |
| ypM | | | | - | | | | - | | | | - |
| MX | 6 (0.12) | 5 (0.15) | 1 (0.07) | | 3 (0.33) | 2 (0.40) | 1 (0.25) | | 8 (0.89) | 5 (0.83) | 3 (1.00) | |
| M0 | 43 (0.88) | 29 (0.85) | 14 (0.93) | | 6 (0.67) | 3 (0.60) | 3 (0.75) | | 1 (0.11) | 1 (0.17) | 0 (0.00) | |

### Table S4. Complete Pre-Therapy MCCV Results for Model Selection

Result format: Mean (SD). Since CLAM pretrained on NSCLC significantly outperforms random initialized CLAM, we did not test it further for other backbones. CLAM pretrained was just examined with ResNet50 encoders because the input feature dimensions need to be $n_{\text{embd}} = 1024$. ViT-4096 was not used for PatchGCN because the node number would be too small and not for CLAM since there are too less regions per patient to aggregate.
Abbreviations: AUROC, area under the receiver operating characteristic curve; AUPRC, area under the precision-recall curve; B. Acc., balanced accuracy; MCC, Matthews correlation coefficient; CLAM pretrained, CLAM network pretrained on TCGA NSCLC and finetuned on MEMORI data

| Decoder | Encoder | AUROC | AUPRC | B.Acc. | MCC |
|---|---|---|---|---|---|
| Max | ResNet50 ImageNet | 0.56 (0.11) | 0.50 (0.08) | 0.51 (0.05) | 0.02 (0.12) |
| | ResNet50 SimTriplet | 0.56 (0.11) | 0.52 (0.12) | 0.51 (0.09) | -0.01 (0.22) |
| | ViT-256 | 0.50 (0.19) | 0.50 (0.15) | 0.50 (0.12) | 0.00 (0.30) |
| | ViT-4k | 0.54 (0.17) | 0.40 (0.11) | 0.52 (0.10) | 0.02 (0.26) |
| Mean | ResNet50 ImageNet | 0.47 (0.16) | 0.45 (0.13) | 0.50 (0.00) | 0.00 (0.00) |
| | ResNet50 SimTriplet | 0.51 (0.09) | 0.47 (0.10) | 0.49 (0.06) | -0.04 (0.16) |
| | ViT-256 | 0.56 (0.21) | 0.57 (0.16) | 0.57 (0.13) | 0.17 (0.28) |
| | ViT-4k | 0.72 (0.12) | 0.67 (0.17) | 0.55 (0.10) | 0.00 (0.00) |
| CLAM | ResNet50 ImageNet | 0.46 (0.13) | 0.45 (0.10) | 0.50 (0.00) | 0.00 (0.00) |
| | ViT-256 | 0.57 (0.15) | 0.54 (0.13) | 0.53 (0.13) | 0.07 (0.30) |
| CLAM-Pretrained | ResNet50 ImageNet | 0.80 (0.14) | 0.76 (0.14) | 0.72 (0.09) | 0.44 (0.18) |
| | ResNet50 SimTriplet | 0.75 (0.15) | 0.69 (0.16) | 0.64 (0.15) | 0.28 (0.30) |
| PatchGCN | ResNet50 ImageNet | 0.72 (0.20) | 0.66 (0.18) | 0.50 (0.00) | 0.00 (0.00) |
| | ResNet50 SimTriplet | 0.74 (0.18) | 0.68 (0.16) | 0.50 (0.00) | 0.00 (0.00) |
| | ViT-256 | 0.58 (0.20) | 0.58 (0.19) | 0.56 (0.09) | 0.14 (0.21) |
